# Healthcare Professionals’ Perspectives on Schizophrenia Recovery: A Literature Review

**DOI:** 10.1101/2025.09.16.25335872

**Authors:** Fandro Armando Tasijawa, Ah Yusuf, Moses Glorino Rumambo Pandin, Joan Herly Herwawan, Violin Irene Ninef, Widya Addiarto, Yade Kurnia Sari, Erika Martining Wardani

## Abstract

**Background:** Recovery is a key concept in mental health care and contributes to improving clinical practice and the quality of life of individuals living with schizophrenia. However, mental health professionals do not yet share a unified perspective and understanding of the recovery process. The aim of this study was to review healthcare professionals’ perspectives on schizophrenia recovery.

**Methods:** A literature search was conducted using electronic databases including PubMed, CINAHL (EBSCO), ProQuest, and ScienceDirect, covering the years 2009 to 2019. The search employed the following keyword combinations: *“Perceptions” AND “Health Workers” OR “Nursing” OR “Doctor” OR “Psychiatrists” AND “Recovery” AND “Schizophrenia*.*”* Eligible studies included original articles with healthcare professionals as participants that examined perspectives on schizophrenia recovery.

**Results:** Out of 439 studies screened, eight met the inclusion criteria: four qualitative studies (n = 79), three quantitative studies (n = 756), and one mixed-methods study (n = 174). Two major themes emerged: positive and negative perspectives. Positive perspectives emphasized that patients could achieve full recovery without long-term medication, engage in daily activities and employment, and experience reduced symptoms. In contrast, negative perspectives conceptualized recovery within a biomedical framework, highlighting barriers such as the impossibility of full recovery, medication dependence, and relapse.

**Conclusion:** Healthcare professionals hold diverse perspectives on schizophrenia recovery. These varying viewpoints may influence both the recovery process of individuals with schizophrenia and decision-making in mental health services. Findings from this review may serve as guidance for healthcare professionals to enhance their understanding of schizophrenia recovery.

## Background

Globally, the recovery-oriented model has been adopted as a national mental health policy in most high-income countries, including the United Kingdom, Wales, and across the European Union (Health, 2011). In contrast, in many low- and middle-income countries, recovery remains relatively new and is still limited in practice, particularly among healthcare providers such as nurses. In these contexts, recovery has not yet become a priority in mental health care, even though individuals living with schizophrenia are in urgent need of recovery-oriented services to support long-term, meaningful, and fulfilling lives.

Recovery was originally defined by Anthony (1993) as “a deeply personal, unique process of changing one’s attitudes, values, feelings, goals, skills, and roles in order to live a satisfying, hopeful, and contributing life—even with the limitations caused by illness.” The contribution of healthcare professionals is critical in shifting mindsets through evidence-based practices that emphasize recovery not as a cure, but as an ongoing journey and a way of life that enables individuals to find meaning through work, social relationships, and community participation (Morrison et al., 2014; Shepherd, Boardman, Rinaldi, & Roberts, 2014; Suryani, 2014). As such, healthcare professionals serve as essential sources of support for individuals with schizophrenia, and their attitudes can profoundly shape how these individuals perceive themselves.

The perspectives of mental health professionals on recovery represent a key component in facilitating the recovery process for people with schizophrenia. Recovery contributes significantly to improvements in mental health practices and to enhancing the quality of life for those diagnosed with schizophrenia. Its implementation in mental health practice requires collaboration with service users, as recovery cannot be fully achieved without valuing their subjective experiences and recognizing their capacity for self-directed recovery (Drapalski et al., 2012). In this context, healthcare professionals do not act as primary agents of change but rather as facilitators—providing resources, encouragement, and opportunities that empower individuals to take responsibility, build confidence, and believe in their ability to recover (Suryani, 2014).

Despite these insights, mental health professionals still hold diverse and sometimes conflicting views on the recovery process. Moreover, to date, no comprehensive review has synthesized their perspectives on recovery in the context of schizophrenia. Understanding these viewpoints is crucial, as they directly influence the resources, motivation, support, quality, and outcomes of recovery-oriented care for individuals living with schizophrenia.

### Objective

The objective of this literature review was to examine healthcare professionals’ perspectives on schizophrenia recovery.

## Methods

### Search Strategy

To minimize the risk of publication bias, a systematic computer-based search was conducted in October 2019 across four electronic databases: PubMed, CINAHL (EBSCO), ProQuest, and ScienceDirect. An advanced search was carried out within the publication period of 2009– 2019. The following keyword combinations were used: *“Perceptions” AND “Health Workers” OR “Nursing” OR “Doctor” OR “Psychiatrists” AND “Recovery” AND “Schizophrenia*.*”*

### Inclusion Criteria

Studies were eligible for inclusion if they investigated healthcare professionals’ perspectives on recovery in psychosis, particularly schizophrenia. Only original research articles employing either qualitative or quantitative methods were considered. Studies from all countries were included; however, non-English publications were excluded from this review.

**Figure 1.**
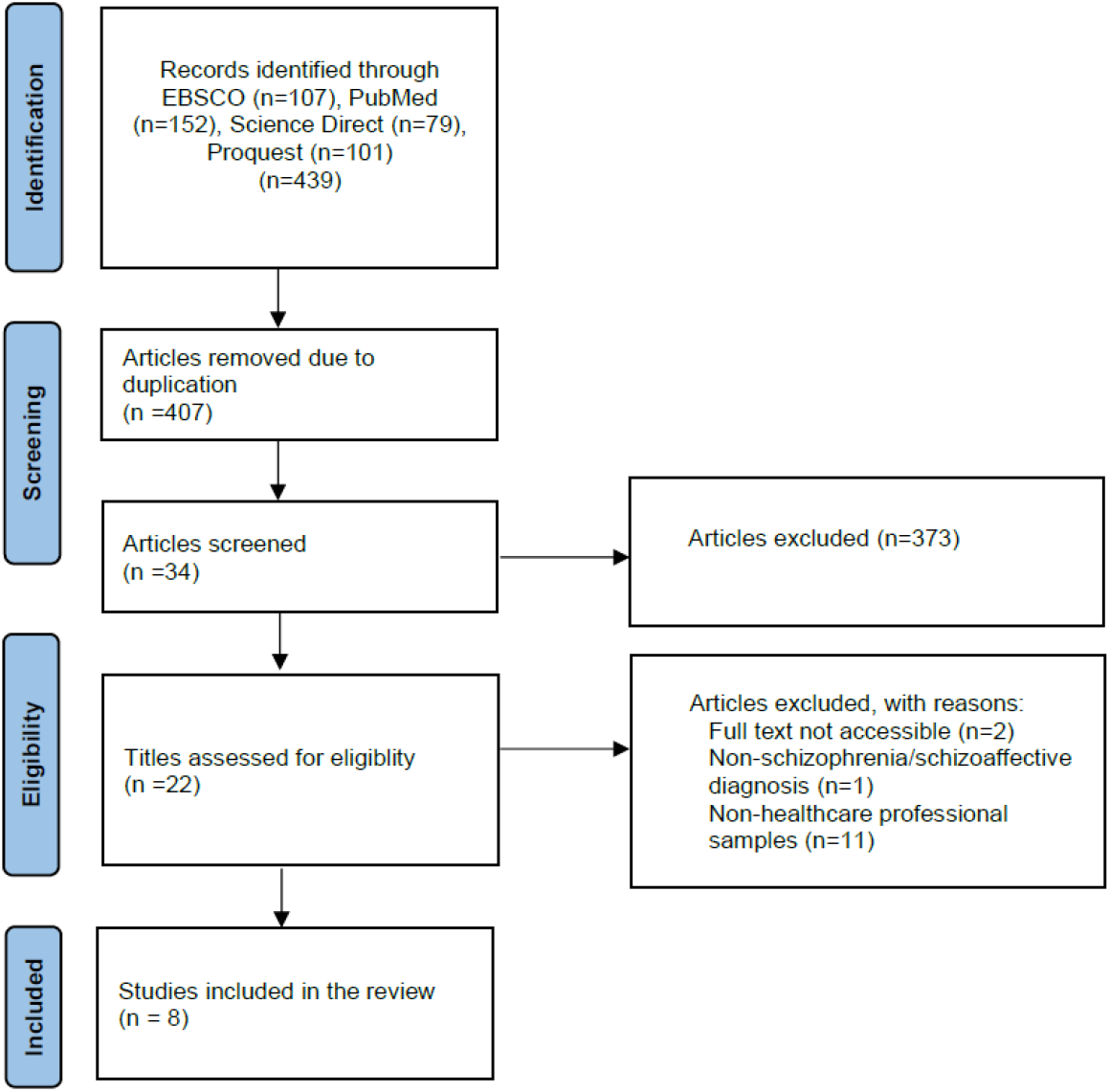
Flowchart of the systematic review selection process

## Results

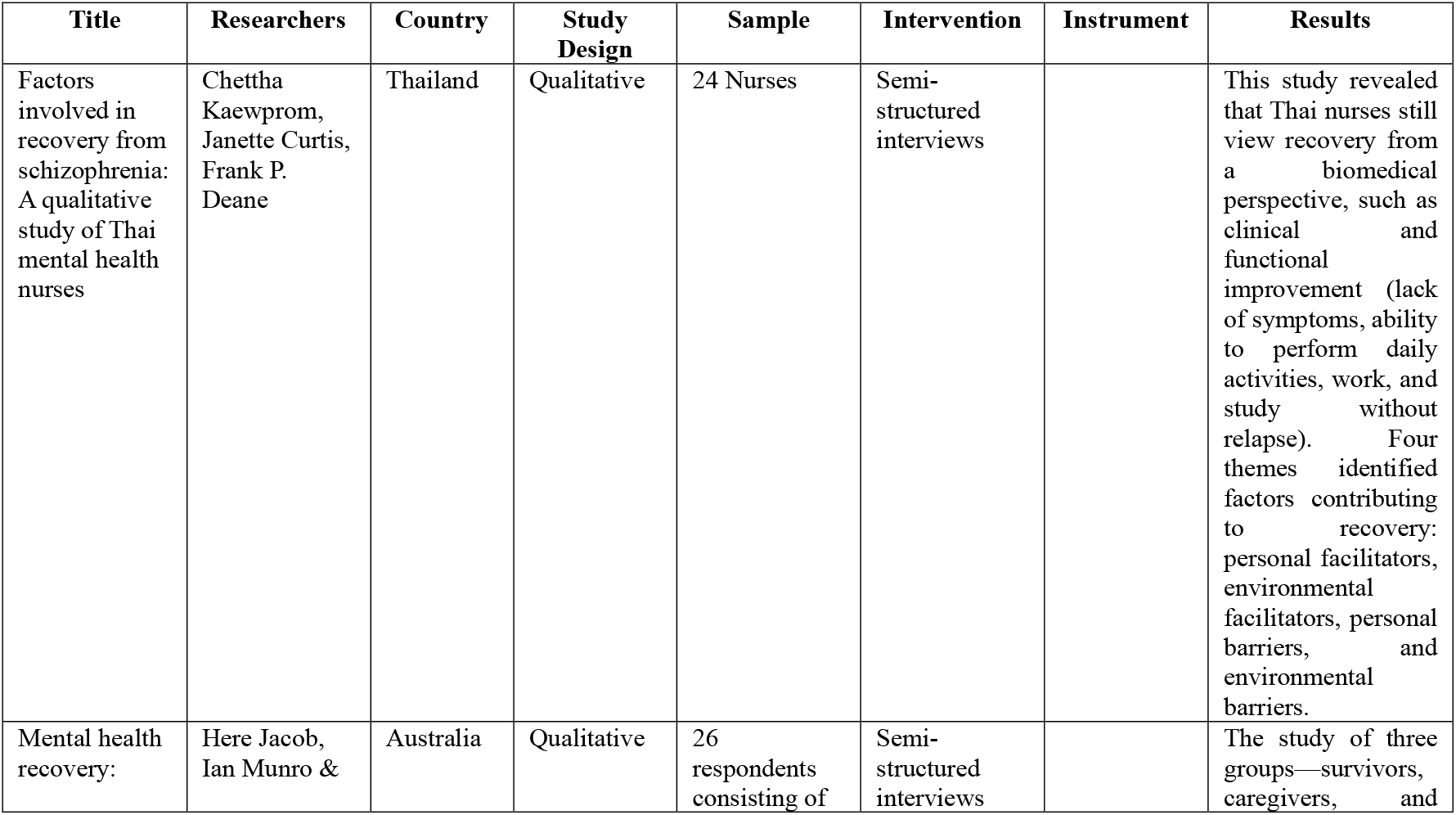
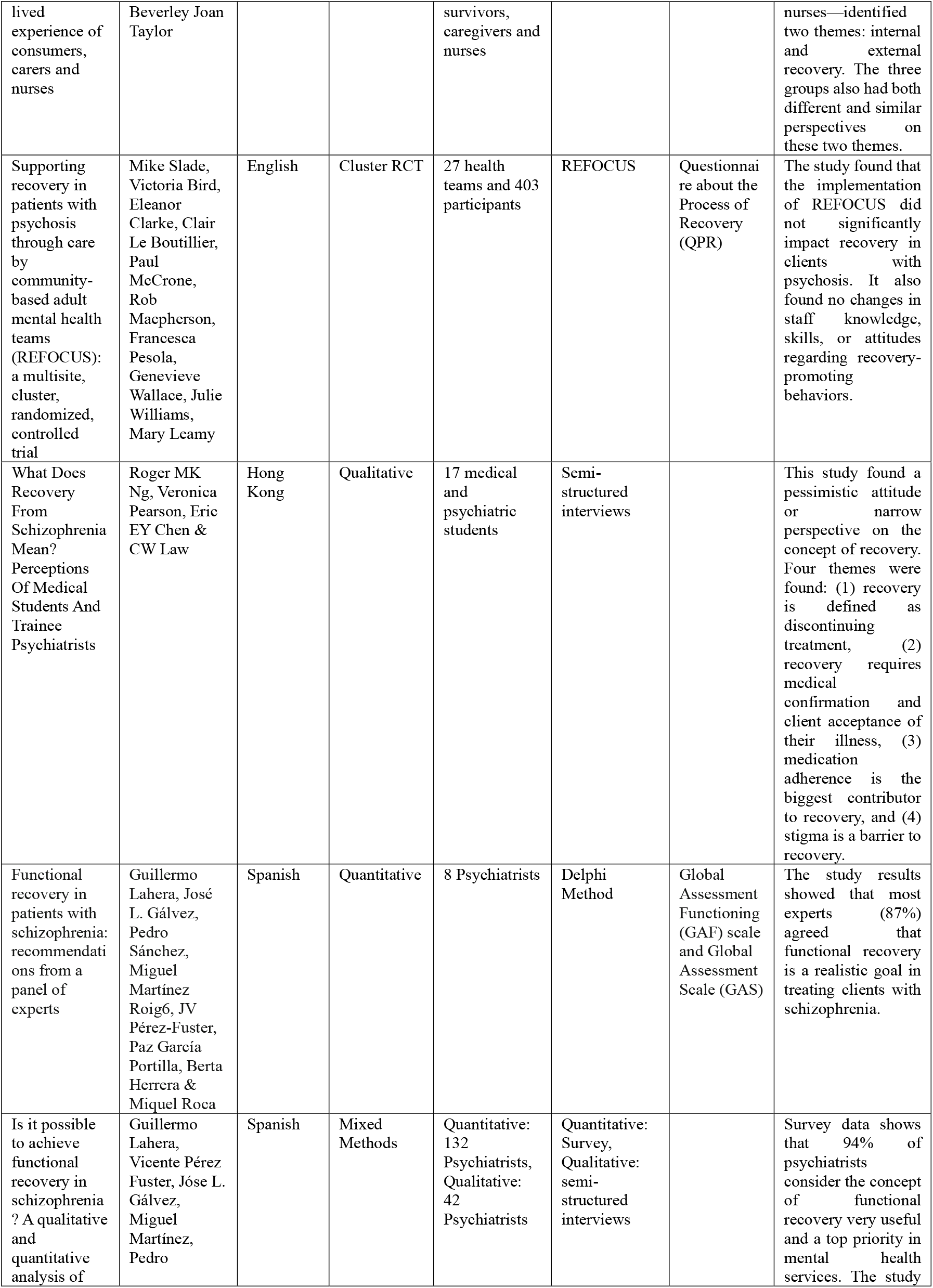
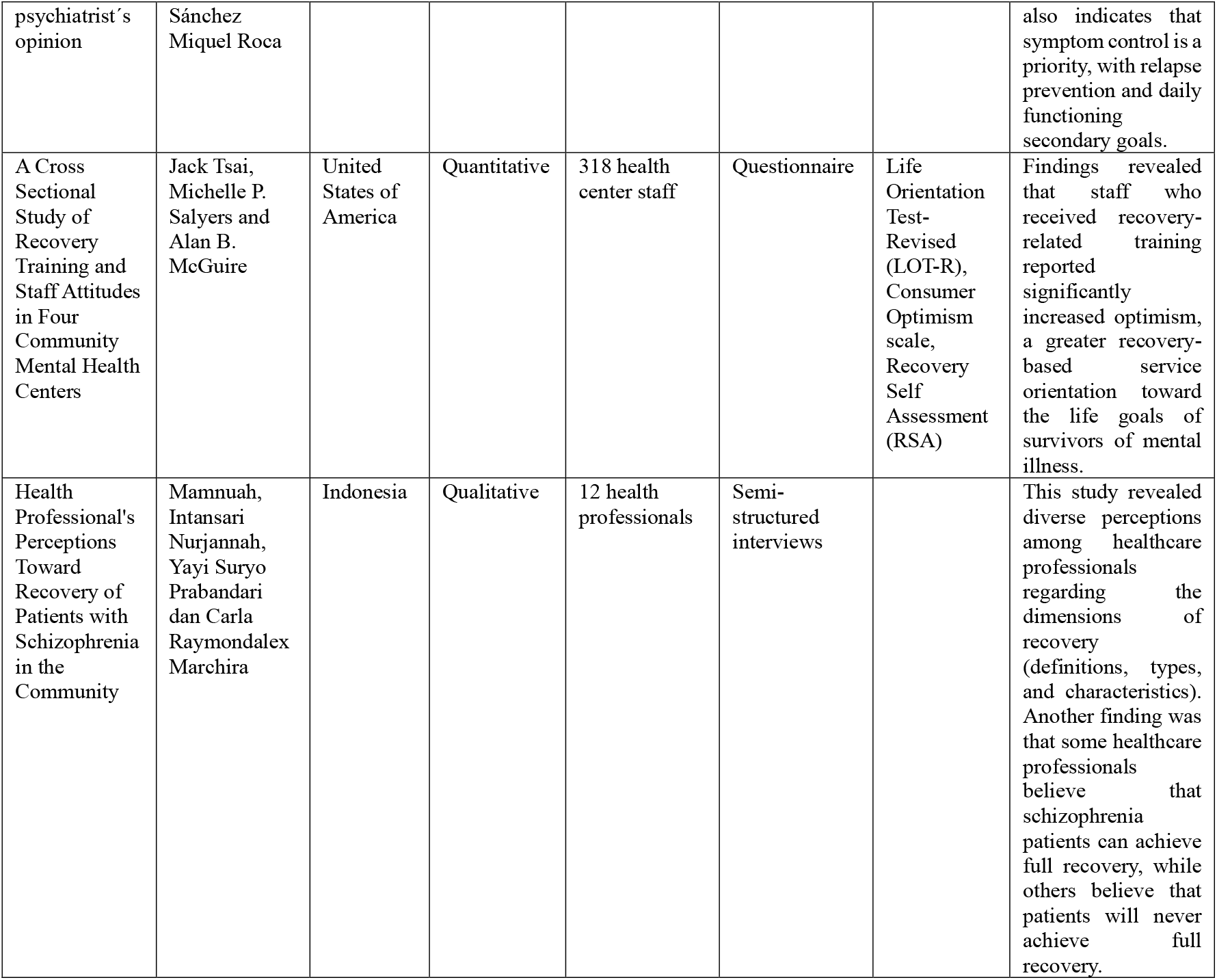

A total of eight studies met the inclusion criteria, consisting of four qualitative studies, three quantitative studies, and one mixed-methods study. These studies were conducted across diverse countries including Thailand, Australia, the United Kingdom, Hong Kong, Spain, the United States, and Indonesia. The participants comprised nurses, psychiatrists, medical students, community mental health staff, and other healthcare professionals.

In Thailand, Kaewprom et al. (2011) found that nurses primarily viewed recovery through a biomedical lens, equating it with clinical and functional improvement such as symptom reduction, the ability to perform daily activities, employment, and education without relapse. They identified four themes influencing recovery: personal facilitators, environmental facilitators, personal barriers, and environmental barriers.

Jacob, Munro, and Taylor (2015) in Australia reported perspectives from service users, caregivers, and nurses. Their findings revealed two overarching themes—internal and external recovery—highlighting both shared and divergent viewpoints across groups.

In the United Kingdom, the REFOCUS trial (Slade et al., 2011) tested the impact of a recovery-oriented intervention among community-based mental health teams. The study found no significant effect on client recovery outcomes, nor changes in staff knowledge, skills, or attitudes toward promoting recovery.

Ng et al. (2011) in Hong Kong explored the views of medical students and trainee psychiatrists, revealing generally pessimistic or narrow understandings of recovery. Four themes emerged: (1) recovery defined as discontinuation of treatment, (2) the need for medical confirmation and patient acceptance of illness, (3) medication adherence as central to recovery, and (4) stigma as a major barrier.

Two studies from Spain (Lahera et al., 2013; Lahera et al., 2014) highlighted psychiatrists’ perceptions. A Delphi panel (n = 8) emphasized that functional recovery was a realistic treatment goal, while a mixed-methods study involving 174 psychiatrists found that 94% regarded functional recovery as both useful and a primary goal in schizophrenia care. Nonetheless, symptom control remained the top priority, with relapse prevention and daily functioning considered secondary.

In the United States, Tsai, Salyers, and McGuire (2010) surveyed 318 community mental health staff. Results showed that recovery training significantly increased staff optimism and orientation toward recovery-oriented service delivery.

Finally, Mamnuah et al. (2017) in Indonesia demonstrated a wide range of healthcare professionals’ perceptions of recovery, particularly regarding its definition, types, and characteristics. Some professionals believed that individuals with schizophrenia could achieve full recovery, while others maintained that full recovery was unattainable.

## Discussion

This review aimed to identify healthcare professionals’ perspectives on schizophrenia recovery. It synthesized findings from both qualitative and quantitative studies, offering complementary insights. Of the eight studies included, four were qualitative (n = 79), three were quantitative (n = 756), and one employed a mixed-methods design (n = 174). A notable strength of this review lies in its diverse study settings, spanning both high-income and low- to middle-income countries, including Thailand, Australia, the United Kingdom, Spain, Hong Kong, the United States, and Indonesia.

The perspectives of healthcare professionals across these countries varied considerably. In Indonesia, Nurjannah, Suryo Prabandari, and Marchira (2019) identified four themes— definitions, types, characteristics, and divergent perceptions of recovery among health professionals. Their qualitative study involving 12 respondents (psychiatrists, psychologists, nurses, primary care doctors, social workers, and provincial health officers) highlighted both positive and negative views. Positive perspectives emphasized the possibility of full recovery without medication, whereas negative perspectives reflected skepticism, assuming permanent dependence on medication and no prospect of complete recovery.

Similar findings were observed in Hong Kong, where Ng, Pearson, Chen, and Law (2011) examined the views of medical students and trainee psychiatrists. Their study revealed narrow and pessimistic perceptions of recovery, identifying four themes: recovery as treatment discontinuation, the primacy of medication adherence, the necessity of medical confirmation and patient acceptance, and stigma as a major barrier. Likewise, Kaewprom, Curtis, and Deane (2011) reported that Thai nurses continued to view recovery through a biomedical lens, focusing on symptom reduction, the ability to resume daily activities, and returning to work or study. Relapse was regarded not as part of the recovery process but as an obstacle, reflecting an incomplete understanding of recovery as a dynamic, non-linear journey.

Contrasting perspectives emerged in Australia. Jacob, Munro, and Taylor (2015), who explored the views of service users, caregivers, and nurses, identified two broad themes—internal and external recovery. Nurses associated recovery with self-acceptance, meaningful living, and regaining pre-illness functioning, even in the presence of symptoms. Meanwhile, psychiatrists in Spain, as reported by Lahera et al. (2018), prioritized symptom control as central to recovery, with relapse prevention and daily functioning as secondary goals. Nevertheless, most psychiatrists agreed that functional recovery was highly valuable, a realistic objective, and a primary aim of schizophrenia care (Lahera et al., 2018; Roca, 2016).

Training was also shown to influence perspectives. A study of 318 community mental health staff in the United States revealed that recovery-focused training was significantly associated with greater optimism and stronger alignment with recovery-oriented care goals compared to staff without such training (Tsai, Salyers, & McGuire, 2011). However, findings by Slade et al. (2015), using the REFOCUS intervention at a team level, reported no significant impact on patient recovery outcomes, nor any change in staff knowledge, skills, or recovery-promoting behaviors.

This review also highlighted several limitations. The majority of included studies were qualitative, providing rich subjective data but limiting generalizability due to small sample sizes. Cultural and ethnic variations further complicate the transferability of findings, as the concept of recovery may be understood differently across contexts. Moreover, disparities in mental health systems—particularly between high-income countries (e.g., the United States, Spain, Australia, Hong Kong) and low- to middle-income countries (e.g., Indonesia, Thailand)—affect the implementation and prioritization of recovery-oriented practices.

## Conclusion

Healthcare professionals’ perspectives on schizophrenia recovery reflect a spectrum ranging from positive to negative. Positive perspectives view patients as capable of achieving full recovery, engaging in meaningful activities, working, and experiencing symptom reduction. Conversely, negative perspectives emphasize a biomedical stance, highlighting medication dependence, relapse, and the perceived impossibility of full recovery. These diverse perspectives significantly influence both the recovery journey of individuals with schizophrenia and decision-making in mental health care services. The findings of this review provide valuable guidance for health professionals to deepen their understanding of recovery-oriented care. Future research should explore these perspectives further, particularly in low- and middle-income countries, focusing on mental health nurses as key stakeholders in community-based services.

## Data Availability

All data supporting the findings of this study are contained within the manuscript.

## Conflict of Interests Statement

The authors declare that there is no conflict of interest regarding the publication of this paper.

## Ethical Approval and Informed Consent Statements

This study did not involve human participants, animals, or patient data; therefore, ethical approval and informed consent were not required.

## Notes

### Competing Interest Statement

The authors have declared no competing interest.

### Funding Statement

This study was funded by the Indonesia Endowment Fund for Education (LPDP), Ministry of Finance of the Republic of Indonesia, under Grant Number: SKPB3682/LPDP/LPDP.3/2025

